# SARS-CoV-2 neutralising antibody activity in a highly vaccinated population: Longitudinal serology studies in Singapore

**DOI:** 10.1101/2022.05.29.22275748

**Authors:** Hannah E Clapham, Wanni Chia, Jinyan Zhang, Lin-Fa Wang, Clarence C Tam

**Author notes:** Author for correspondence: Dr Clarence C Tam, Saw Swee Hock School of Public Health, National University of Singapore, Tahir Foundation Building, 12 Science Drive 2, Singapore 117549, Singapore.

## Abstract

**Background:** There is continuing uncertainty regarding the longevity of immunological responses to both SARS-CoV-2 natural infection and COVID-19 vaccines.

**Methods:** We analysed data from two serological cohorts in Singapore among residents of a COVID-19 affected migrant worker dormitory between May-July 2020, and among mRNA COVID-19 vaccine recipients between May 2021 and January 2022. We compared SARS-CoV-2 neutralising antibody levels by age group, sex, presence of pre-existing medical conditions, type of mRNA vaccine received and number of doses received. We investigated the effect of time since infection or vaccination on antibody levels in naturally infected individuals and two- and three-dose vaccinees.

**Results:** After two vaccine doses, neutralising antibody responses were higher in Spikevax (Moderna) recipients, females, younger individuals and those with no underlying medical conditions. However, antibody levels waned to similar levels in all groups over time. A third dose boosted these to similarly high levels in all groups. Waning was apparent among two-dose but not three-dose recipients over a period of six months. Both two and three-dose vaccine recipients showed consistently higher neutralising antibody levels compared with naturally infected individuals over the 12-week period following infection or vaccination.

**Conclusions:** Our findings support the broad use of booster doses to improve population protection from COVID-19. However, recent increases in transmission of new SARS-CoV-2 variants, even in the presence of high levels of neutralising antibody in a highly vaccinated population, point to vaccine breakthrough as an important mechanism for maintaining SARS-CoV-2 circulation and indicate the need for variant-specific or universal COVID-19 vaccines.

**Summary:** Younger individuals, females and those with no pre-existing conditions have higher neutralising antibody levels after two doses of COVID-19 mRNA vaccine. Subsequently these wane to levels seen in other groups. A booster dose promotes similarly high levels in all groups.

## Introduction

Singapore is a high-income, island city state in the tropics. The first case of COVID-19 was identified on January 23^rd^, 2020. Despite intensive control measures, including testing, contact tracing, isolation and quarantine, and community-wide social distancing and infection prevention and control, the country experienced explosive epidemics in migrant worker dormitories in mid-2020. Subsequently, large waves of transmission were seen in late 2021 and early 2022 associated with the Delta and Omicron SARS-CoV-2 variants.

The national COVID-19 vaccination programme began in January 2021, initially prioritising the elderly, those with pre-existing medical conditions, and healthcare and essential workers, before being extended to younger adults, adolescents and children aged >5 years. Comirnaty (Pfizer-BioNTech) and Spikevax (Moderna) are the main vaccines used, with inactivated vaccines (Sinovac-Coronavac and Sinopharm) available for those ineligible for mRNA vaccines. Singapore is currently one of the most highly vaccinated populations, with >90% of the population having received a COVID-19 primary vaccination series and >70% having received a booster dose as of April 2022.

Between May and July 2020, we conducted a longitudinal study to investigate infection rates and immunological responses to SARS-CoV-2 infection among migrant workers over a six-week period. Subsequently, we initiated a longitudinal, community-based SARS-CoV-2 serology study in November 2020, with follow-ups after six and twelve months. In this paper, we report on the serological responses of mRNA vaccine recipients over time and compare them with serological responses observed after natural infection.

## Methods

The characteristics of the two cohorts are described in detail elsewhere.[1] The migrant worker cohort included 541 adult males aged 19-59 years residing in a COVID-19 affected dormitory and who provided blood samples in May 2020 and subsequently after two and six weeks. The community cohort comprised 937 community-dwelling adults aged ≥21 years who provided blood samples for serological analysis in November/December 2020 (Figure 1). Of these, 890 (94.9%) provided a subsequent blood sample in May/June 2021 and 868 (92.6%) in January 2022. We analysed samples for SARS-CoV-2 neutralising activity using a surrogate Virus Neutralisation Test (sVNT) (cPass, GenScript).[2,3] At each follow-up, participants also provided information on the presence of other medical conditions, including diabetes, hypertension and heart disease, as well as the number of COVID-19 vaccine doses received, the type of vaccine, and the last date of vaccination.

**Figure 1:**
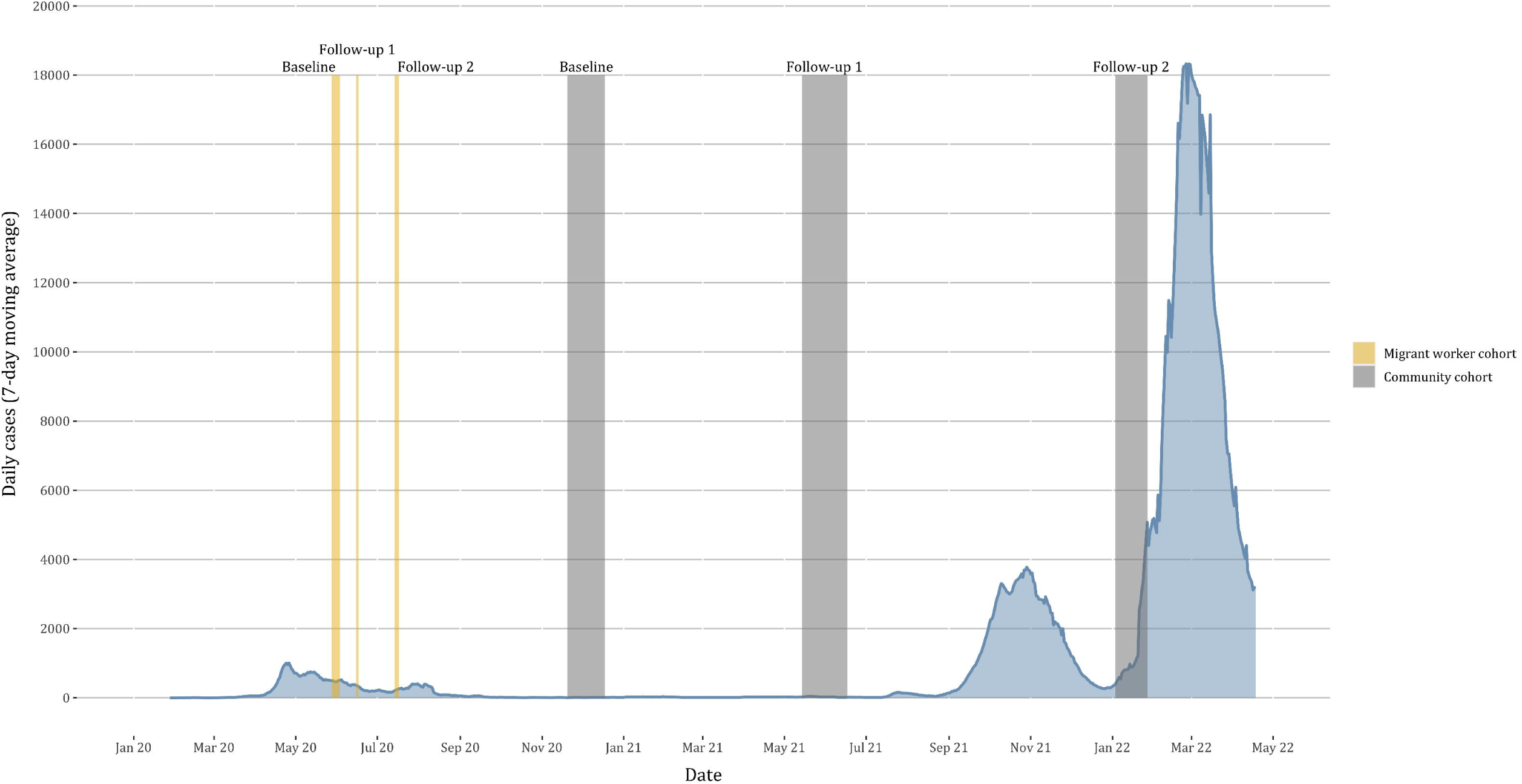
Timeline of COVID-19 epidemic in Singapore and sampling dates in migrant worker and general population cohorts

In this analysis, we included serological samples from 515 participants who had received two doses of a COVID-19 mRNA vaccine by May/June 2021, and samples from 136 and 632 participants who had received two and three doses of vaccine by January 2022. For two- and three-dose vaccine recipients, we compared SARS-CoV-2 neutralising activity by vaccine type (Comirnaty/Spikevax) using Wilcoxon ranksum tests. We then compared cumulative distributions of neutralising activity by time point and number of vaccine doses for different population subgroups defined by sex, age group and pre-existing medical conditions. Finally, we estimated neutralising activity decay rates for two- and three-dose mRNA vaccine recipients by modelling neutralising activity levels by time since receipt of the second or third dose. We used a multivariable Beta regression adjusting for participant sex, age group, vaccine type and pre-existing medical conditions, excluding individuals who had received their last dose <7 days from the date of blood sampling.

Additionally, we compared neutralising antibody activity between mRNA vaccinees from the community cohort and individuals with mild or asymptomatic SARS-CoV-2 infection in the migrant worker cohort, 245 (51.3%) of whom seroconverted between April and July 2020. To maximise comparability between cohorts, we restricted the analysis to males aged 19 to 59 years. We compared neutralising antibody levels by time period since receipt of the second and third dose in the community cohort, or since seroconversion in the migrant worker cohort (<2 weeks, <4 weeks, 4-6 weeks, 6-12 weeks).

Analyses were performed in R version 4.1.[4]

### Ethics statement

The community cohort study was approved by the National University of Singapore institutional review board (reference H-20-032). The migrant worker cohort study was approved by the Singapore Ministry of Health under the Infectious Diseases Act (Schedule 59A) as part of the national public health response to the COVID-19 epidemic.

## Results

Of the 799 individual vaccinees included in this analysis, 553 had received only Comirnaty vaccine, 131 Spikevax only and 115 mixed doses of Comirnaty/Spikevax vaccine. Among two-dose vaccinees, median time to receipt of the second dose was 27 days (interquartile range, IQR: 12.5 – 51.5 days) at the May/June 2021 follow-up and 157 days (IQR: 137.0 – 171.5 days) at the January 2022 follow-up. Median time to receipt of a third dose at the January 2022 follow-up was 64 days (IQR: 36 – 92 days).

Spikevax vaccine recipients had significantly higher neutralising antibody activity levels after two doses (p=0.006) (Figure 2a). These differences were negated in three-dose vaccine recipients, who generally had very high neutralising antibody levels, irrespective of which vaccine(s) they received; single vaccine and mixed dose recipients demonstrated similar neutralising antibody activity (Figure 2b).

**Figure 2:**
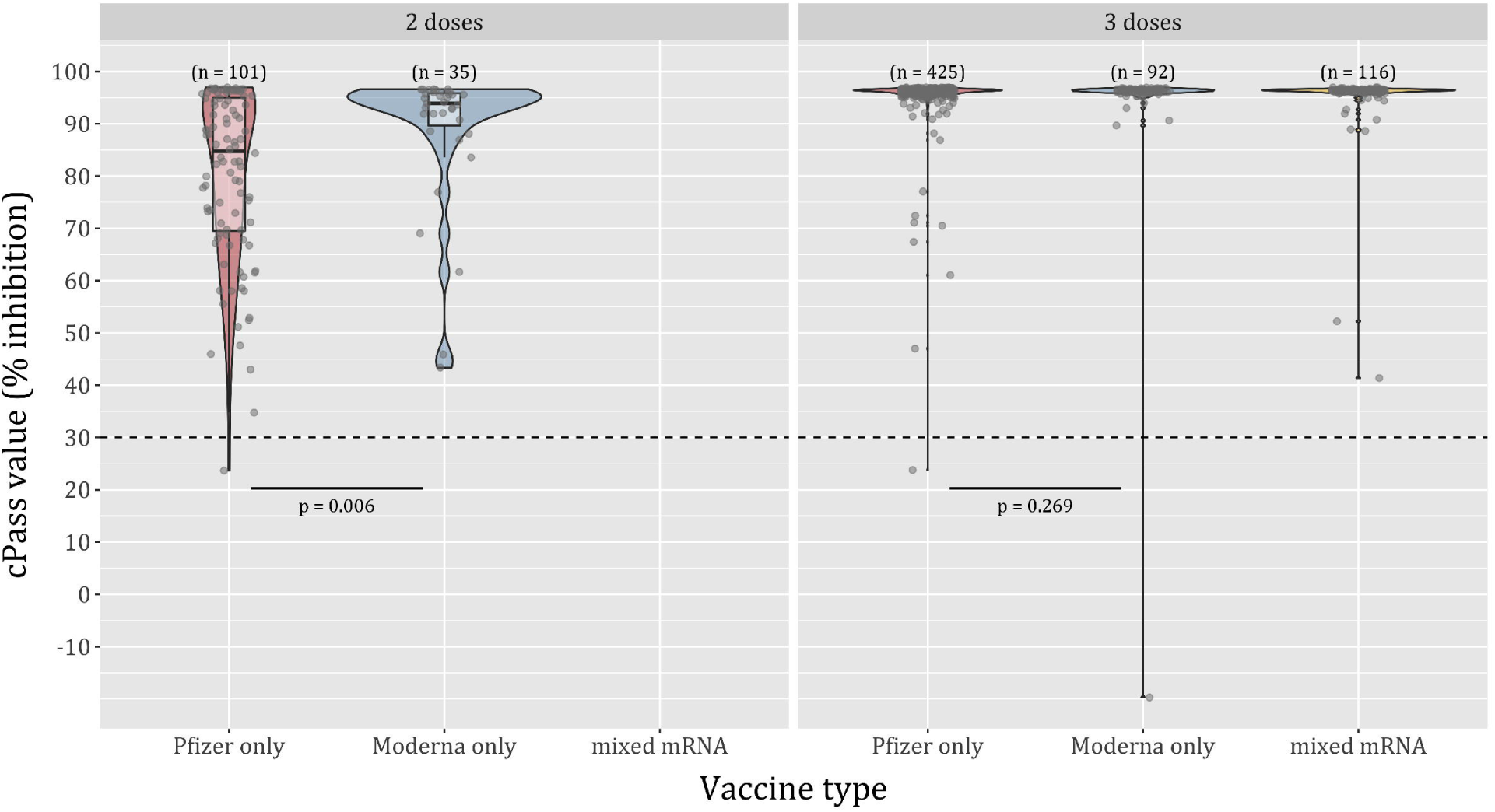
Distribution of SARS-CoV-2 neutralising antibody activity by vaccine type and number of doses. Panel A: 2-dose vaccine recipients; Panel B: 3-dose vaccine recipients

When comparing cumulative distributions, older individuals, particularly those aged ≥70 years, had lower overall neutralising activity responses to two doses in May/June 2021 (Figure 3a). By January 2022, however, all age groups showed lower and similar neutralising antibody levels, indicating that, despite the initial higher responses in younger individuals, these wane to similar levels in all age groups over time (Figure 3b). Among three-dose recipients in January 2022, however, neutralising activity was much higher than in two-dose recipients and similar across age groups (Figure 3c).

**Figure 3:**
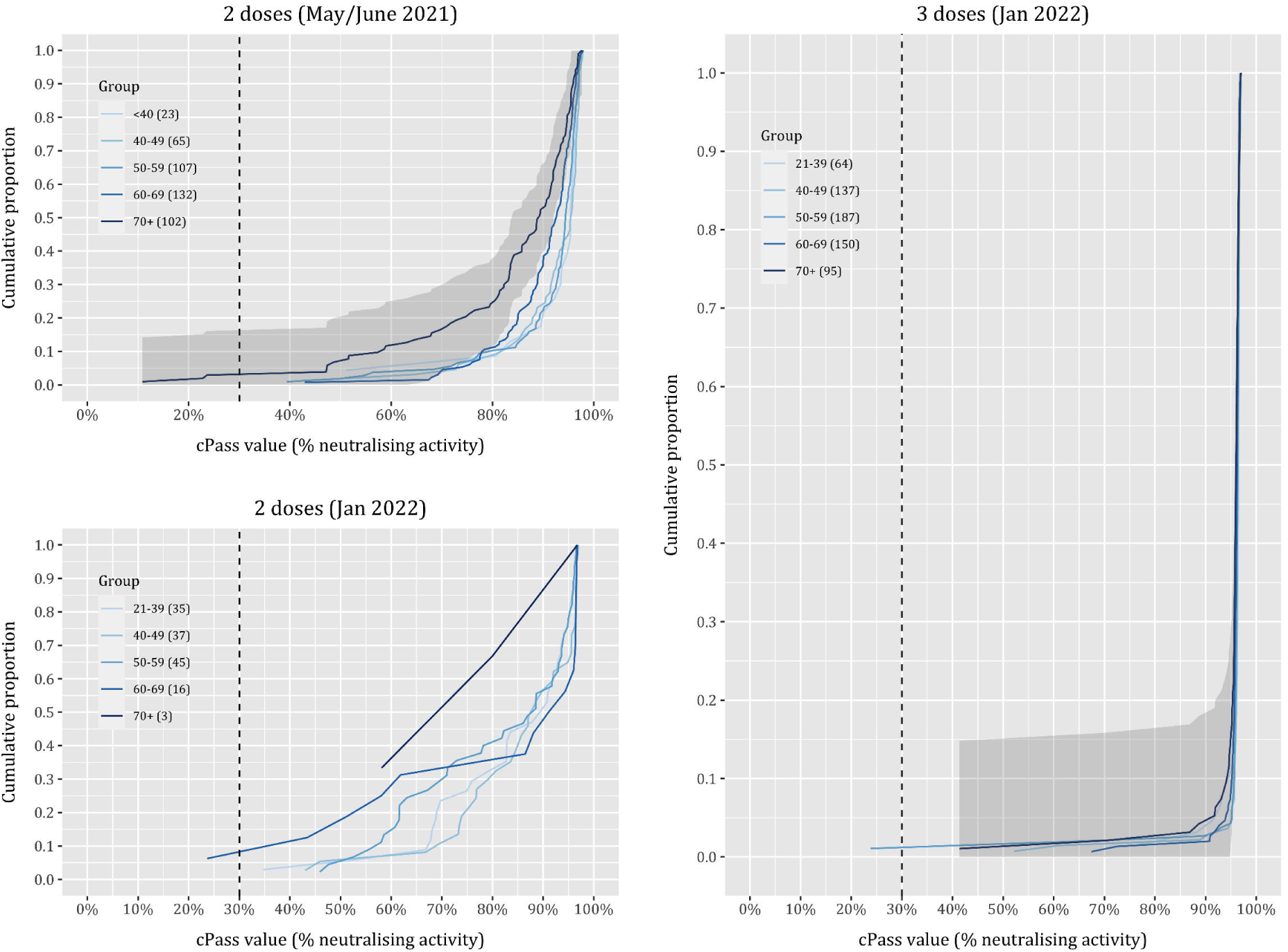
Cumulative distribution of SARS-CoV-2 neutralising antibody activity by age group, timepoint and number of doses. Top left: 2-dose vaccine recipients in May/June 2021; Bottom left: 2-dose recipients in January 2022; Right: 3-dose vaccine recipients in January 2022

We observed similar patterns of differential waning by sex and pre-existing medical conditions. Despite higher initial antibody responses to two doses in May/June 2021 among females (Supplementary Figure S1a), levels waned to similar levels as males by January 2022 (Supplementary Figure S1b). A third dose boosted these to much higher levels with minimal differences between sexes (Supplementary Figure S1c). Similarly, those with diabetes and hypertension had lower initial responses to two doses in May/June 2021 compared with participants with no pre-existing conditions (Supplementary Figure S2a). In January 2022, however, two-dose recipients showed similar neutralising antibody levels irrespective of pre-existing medical conditions (Supplementary Figure S2b), and three-dose recipients had uniformly high neutralising activity (Supplementary Figure S2c).

After adjusting for participant sex, age group, vaccine type and pre-existing medical conditions, waning was apparent among two-dose recipients, but neutralising activity remained high among three-dose recipients even up to more than six months after receipt of the third dose (Figure 4a).

**Figure 4:**
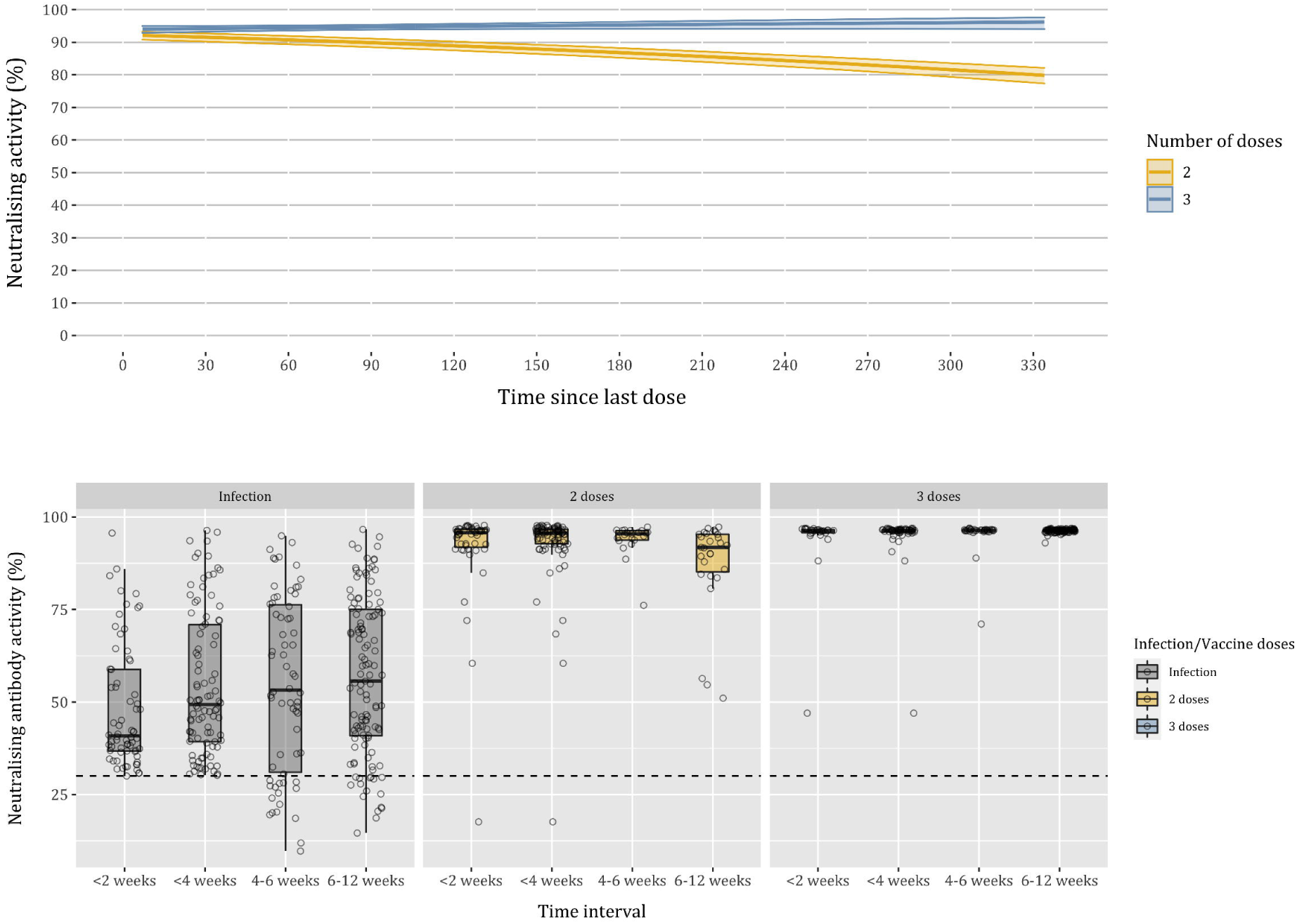
SARS-CoV-2 neutralising antibody activity by time since vaccination or natural infection. Top: SARS-CoV-2 neutralising antibody activity by time since receipt of second or third dose of a COVID-19 mRNA vaccine. Bottom: SARS-CoV-2 neutralising antibody activity in mildly and asymptomatically infected migrant workers (left), two-dose mRNA vaccine recipients (centre), and three-dose mRNA vaccine recipients, by time since infection or vaccination

### Neutralising antibody activity after vaccination and natural infection

When comparing neutralising antibody activity between vaccinees and mild or asymptomatically infected individuals, males aged <60 years in the general population cohort who had received two or three doses within the past 12 weeks showed consistently higher neutralising antibody activity compared with mildly or asymptomatically infected males of the same age group in the migrant worker cohort (Figure 4b).

### Infection risk in unvaccinated individuals

At the January 2022 follow-up there were 51 unvaccinated individuals who were seronegative in May/June 2021. Of these, 43 (84%) had neutralising antibody activity against SARS-CoV-2, indicating a high force of natural infection within the previous 6-7 months.

## Discussion

Our findings provide further insights into the serological response to COVID-19 mRNA vaccines. In particular, we found that although males, older individuals and those with certain pre-existing medical conditions initially mount less potent neutralising antibody responses to two doses of vaccine, neutralising activity nonetheless wanes to similar levels in all groups over time. A third dose has an equalising effect, boosting neutralising antibody activity to similarly high levels in all groups that appear to be maintained even after six months.

We have previously shown that despite large epidemics in migrant worker dormitories, Singapore experienced very low levels of transmission in the wider community during first year of the pandemic, with <0.2% of the population showing serological evidence of SARS-CoV-2 infection.[1] Transmission gradually increased in 2021, resulting in large epidemic waves associated with the highly-transmissible Delta (B.1.617.2) and Omicron (B.1.1.529) variants in late 2021 and early 2022, and coinciding with the easing of travel restrictions and social management measures. The dramatic rise in the SARS-CoV-2 force of infection associated with the Delta variant is reflected in the high levels of seroconversion among unvaccinated individuals between May/June 2021 and January 2022. Despite this, COVID-19 mortality in Singapore has remained comparatively low throughout the pandemic, with a case fatality of <0.3%, in large part because of high levels of vaccine uptake; as of March 2022, 93% of the population has completed a primary course of vaccination and ∼70% has received a booster dose. Taken together, these observations provide support for the widespread use of booster doses regardless of age or pre-existing conditions to ensure universally high neutralising antibody levels and counteract rapid antibody waning over time. At the height of the Omicron wave in early March, two-dose vaccine recipients had a 3.5 times higher risk of being critically ill and intubated in an intensive care unit compared with three-dose vaccinees; the relative risk among unvaccinated individuals was 7.5 times higher.[5]

There are two important caveats. First, the sVNT assay used in this study was developed based on the ancestral SARS-CoV-2 strain; neutralising activity specifically against the Delta and Omicron variants is likely to be lower. This is reflected in the dramatic rises in Delta and Omicron transmission in Singapore in late 2021 and early 2022 despite very high levels of vaccine uptake, which coincide with the easing of travel restrictions and social distancing measures. Second, there is continuing uncertainty regarding the relative protection afforded by previous infection and vaccination, as well as the role of neutralising antibodies in immune protection. A recent systematic review suggested that previous infection provided equivalent protection from COVID-19 compared with two doses of mRNA vaccine,[6] although other studies have shown superior protection from two doses of vaccine.[7,8] Our findings indicate considerably lower neutralising antibody levels following natural infection relative to two and three vaccine doses. A major consideration is that infections in our migrant worker cohort were mild or asymptomatic, which tend to elicit lower, more rapidly waning neutralising antibody responses.[9] Furthermore, cell-mediated responses are known to play an important role in the immune response to SARS-CoV-2. Mild and asymptomatic SARS-CoV-2 infections have been shown to produce robust, durable cell-mediated responses,[10] and early induction of SARS-CoV-2 specific T cells appears to be associated with faster viral clearance.[11] Recent studies have also shown that effective T-cell responses are also maintained against the Delta and Omicron variants.[12] High neutralising antibody titres and robust cell-mediated responses are both likely to be responsible for the dramatic reduction in risk of severe illness and COVID-19 mortality among vaccinated individuals. Despite this, recent increases in SARS-CoV-2 transmission following the emergence of new virus variants, even in the presence of high levels of neutralising antibody in a highly vaccinated population, point to vaccine breakthrough as an important mechanism for maintaining SARS-CoV-2 circulation and indicate the need for the development of variant-specific or universal COVID-19 vaccines.

## Data Availability

All data produced in the present study are available upon reasonable request to the authors

## Acknowledgements

Recruitment and data collection for the cohort study was made possible by the Singapore Population Health Studies operations team.

