## Supplementary information for "SARS-CoV-2 neutralising antibody activity in a highly vaccinated population: Longitudinal serology studies in Singapore"

Figure S1: Cumulative distribution of SARS-CoV-2 neutralising antibody activity in males and females by timepoint and number of doses. Panel A: 2-dose vaccine recipients in May/June 2021; Panel B: 2-dose recipients in January 2022; Panel C: 3-dose vaccine recipients in January 2022

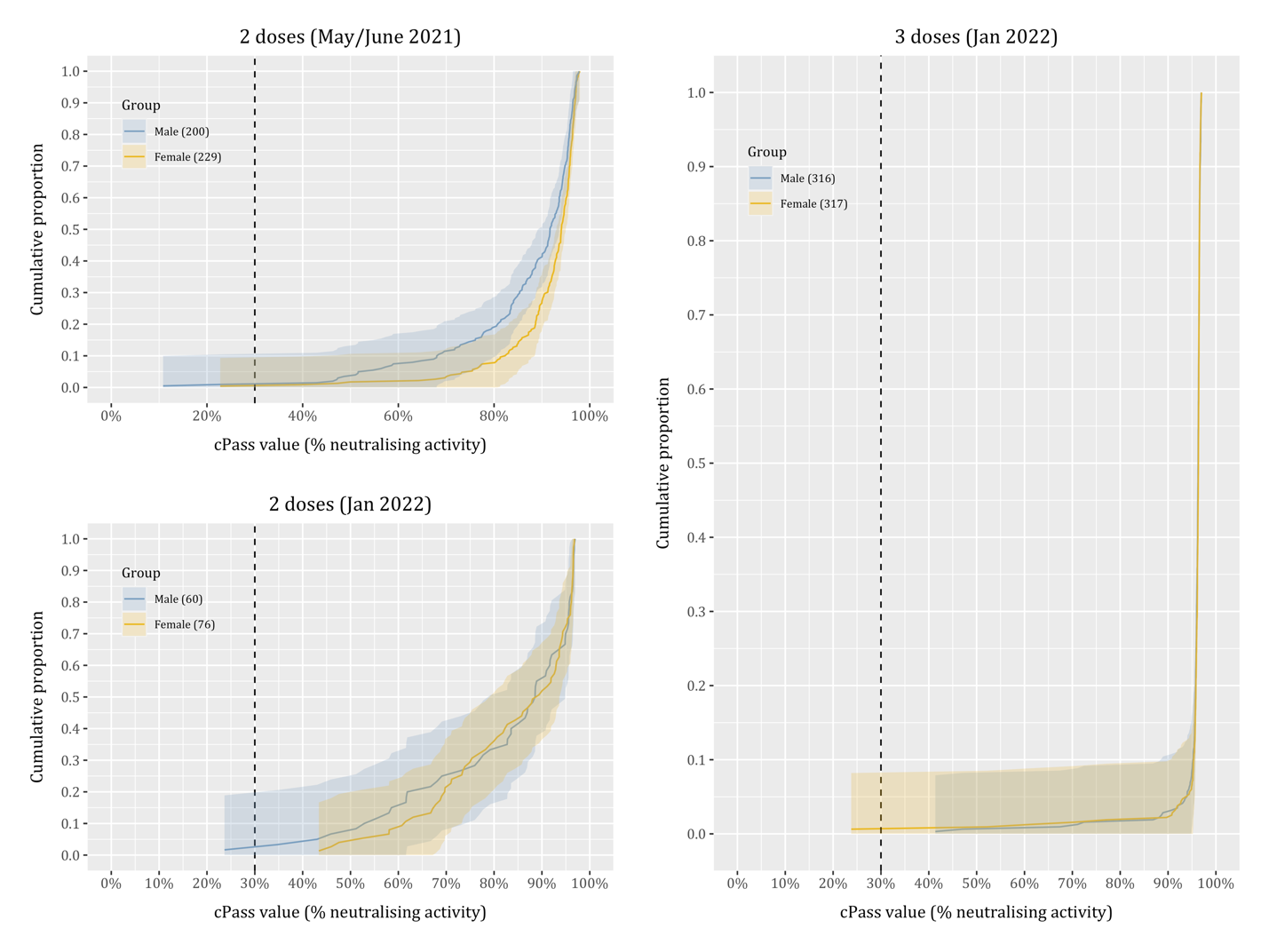

Figure S2: Cumulative distribution of SARS-CoV-2 neutralising antibody activity by pre-existing medical conditions, timepoint and number of doses. Panel A: 2-dose vaccine recipients in May/June 2021; Panel B: 2-dose recipients in January 2022; Panel C: 3-dose vaccine recipients in January 2022

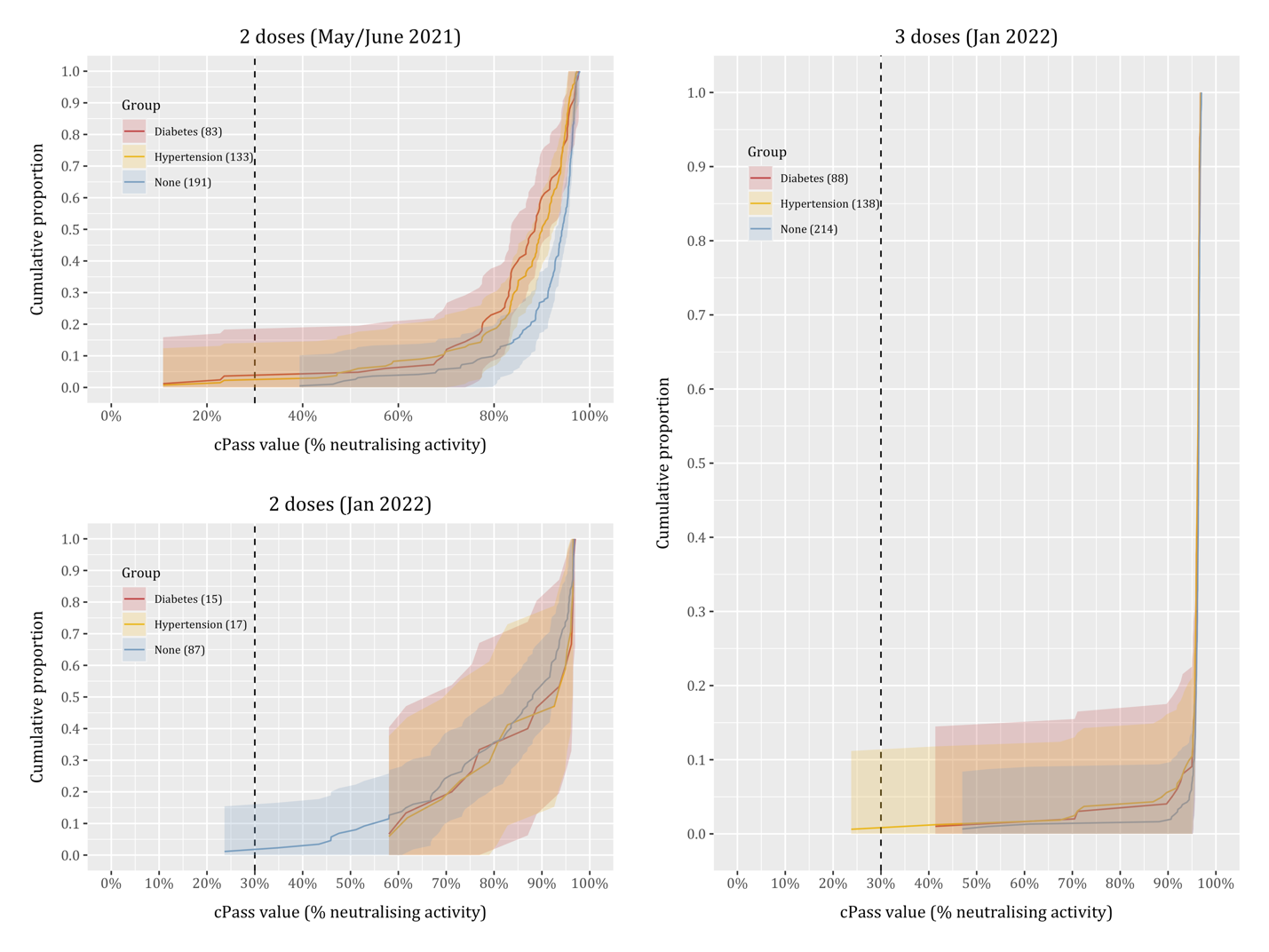
